# Validation of aEEG-CSA Neonatal Seizure Detection Algorithm on Hypothermia Treated Infants with HIE

**DOI:** 10.64898/2026.07.02.26356964

**Authors:** Sylvia Edoigiawerie, Julia Henry, Brett Beaulieu-Jones, Henry David, Naoum P. Issa

## Abstract

**Objective:** To validate a neonatal seizure detection algorithm that is based on extracted clinical features of the aEEG and CSA on a cohort of cooled neonatal patients with hypoxic ischemic encephalopathy.

**Methods:** A seizure detection algorithm was designed using aEEG margin features, CSA features, trained on a public dataset of 79 neonatal EEGs with three supervised machine learning classifiers. It was subsequently tested on an inhouse cohort of 23 neonates with asphyxia whose EEGs were collected during cooling therapy.

**Results:** The trained Random Forest, Support Vector Machines and Artificial Neural Network classifiers had an AUC of 0.76, 0.77, and 0.77 and an average accuracy of 0.85, 0.86, and 0.85 respectively. Finally, the average AUC across the 10 seizure patients included was 0.85.

**Conclusion:** A neonatal seizure detection algorithm that uses a combination of aEEG and CSA clinical features can capture seizures in HIE patients. Performance across seizure patients is not correlated with seizure duration.

## Introduction

Hypoxic Ischemic Encephalopathy (HIE) is the primary cause of neonatal seizures (1). HIE is not only responsible for 60 to 80% of neonatal seizures, but it also represents a major global disease burden since birth asphyxia is associated with about a quarter of all neonatal deaths (1–3). Therapeutic hypothermia, or cooling therapy, is the standard treatment for moderate to severe HIE. This procedure involves lowering the infant’s core temperature from 98.6 to 92.3 degrees Fahrenheit for 72 hours to reduce secondary energy failure and limit ongoing neuronal injury (4). During cooling therapy, electroencephalography (EEG) is necessary to identify and treat neonatal seizures, but because EEG interpretation is time consuming, it slows down treatment. Seizure burden is particularly important to capture accurately during cooling therapy because high seizure burden has been associated with poor neurocognitive outcome, and infants’ brains are most vulnerable to seizures within the first few days of life (5,6).

Neonatal seizures are also far more difficult to detect than adult seizures. Adult seizures typically produce clear motor or behavioral changes, but neonatal seizures are often subtle or entirely electrographic because neonatal cortical networks are immature and poorly myelinated (7). As a result, more than half of neonatal seizures have no observable clinical correlate (8), making EEG the only reliable detection method (9). Since EEG is resource intensive and time consuming, clinicians are increasingly turning towards quantitative electroencephalography (qEEG) tools to expedite seizure detection. Thus, there is a need for sensitive and specific seizure detection algorithms to expedite clinical management and potentially reduce the morbidity and mortality associated with prolonged and undetected seizures. The primary challenge to generating neonatal seizure detection algorithms is limited availability of annotated neonatal EEG recordings for training and testing. This limitation exists because of the extensive expertise required to identify neonatal seizures on EEG and the low supply of pediatric neurologists with this expertise. Indeed, the first and to the best of our knowledge the only publicly available dataset of neonatal EEG recordings with multiple expert annotations was released in 2019 from Helsinki University (10).

Currently, the amplitude-integrated EEG (aEEG) is the standard bedside qEEG tool for neonates (11); the Compressed Spectral Array (CSA) is more widely used for adults and has not been exclusively tested for manual seizure detection in a neonate-only cohort. Both tools are used by clinicians to manually find EEG periods that might be suspicious for seizure activity. The standard bedside aEEG shows low sensitivity for manual seizure detection with around 25% of individual seizures detected using aEEG (12). Thus, there is a need for computational approaches that can improve performance.

The objective of this study is to evaluate whether a neonatal seizure detection algorithm trained on the publicly available Helsinki University dataset can generalize to an independent cohort of term neonates with HIE, who underwent therapeutic hypothermia at University of Chicago. Our algorithm extracts clinically meaningful time and frequency domain features from both aEEG and CSA. These features are then used to train supervised machine learning classifiers that form an aEEG-CSA ensemble model. In prior work, this ensemble model was trained and tested solely within the Helsinki dataset, which includes neonates with heterogeneous etiologies. Here, we assess whether a model trained on that external dataset can accurately detect seizures in a separate cohort of cooled HIE neonates, thereby testing its transferability and real-world applicability.

## Methods

### Datasets for Training and Testing

The training dataset was taken from a public dataset of neonatal EEG recordings from Helsinki University (10). This dataset contains 79 patients, 39 of whom had seizures. The dataset was annotated for seizures by three reviewers, and consensus labels were used for training. Based on the consensus annotation, there are a total of 302 seizures in the training set.

For the testing dataset, EEG recordings from a cohort of 23 term neonates with HIE were collected from UChicago’s Comer Children’s hospital (Table 1). All recordings were taken during cooling therapy within the first 48 hours of life and extracted from Natus Neuroworks EEG software. For the testing set, EEG recordings were annotated by a pediatric neurologist (JH) with extensive expertise in neonatal epilepsy and over ten years of experience. Seizure annotations for the testing cohort were also cross-referenced with the neonatal EEG chart report taken during cooling therapy and read by other experienced pediatric neurologists at the UChicago Comer’s children’s hospital. There is a total of 105 seizures in the testing set.

**Table 1:** Gestational age, total seizure burden in minutes, and recording duration per patient.

| *Patient Number* | *Gestational Age (Weeks)* | *Seizure Burden (min)* | *Recording Duration (min)* |
| --- | --- | --- | --- |
| **1** | 40.3 | 10.6 | 45.3 |
| **2** | 35.6 | 46.0 | 60.7 |
| **3** | 40.1 | 0 | 60.1 |
| **4** | 37.0 | 0.97 | 41.9 |
| **5** | 40.0 | 3.40 | 59.8 |
| **6** | 40.3 | 37.6 | 46.5 |
| **7** | 40.0 | 24.2 | 60.4 |
| **8** | 41.0 | 68.6 | 145.8 |
| **9** | 38.6 | 0 | 60.2 |
| **10** | 38.0 | 0 | 60.1 |
| **11** | 40.4 | 60.3 | 239.2 |
| **12** | 38.6 | 0 | 60.1 |
| **13** | 38.7 | 0 | 60.1 |
| **14** | 38.1 | 0 | 60.1 |
| **15** | 39.0 | 0 | 60.1 |
| **16** | 40.4 | 0 | 60.1 |
| **17** | 39.9 | 0 | 60.1 |
| **18** | 38.9 | 0 | 60.1 |
| **19** | 38.4 | 0 | 60.1 |
| **20** | 41.1 | 26.5 | 193.5 |
| **21** | 39.3 | 0 | 60.0 |
| **22** | 39.1 | 96.9 | 367.1 |
| **23** | 40.4 | 0 | 60.1 |

### EEG Preprocessing and aEEG-CSA Neonatal Seizure Detection Algorithm

All EEG recordings were preprocessed using bandpass filtering from 0.5 to 40 Hz using a sixth-order Butterworth filter and the MATLAB function “filtfilt” to prevent phase distortion. EEGs were placed in a bipolar montage using centroparietal electrodes C3-P3 and C4-P4 (Figure 1A). These electrodes are most sensitive for capturing neonatal seizures (12–14). Computations and data analysis were done using MATLAB 2022b and GraphPad Prism.

**Figure 1.**
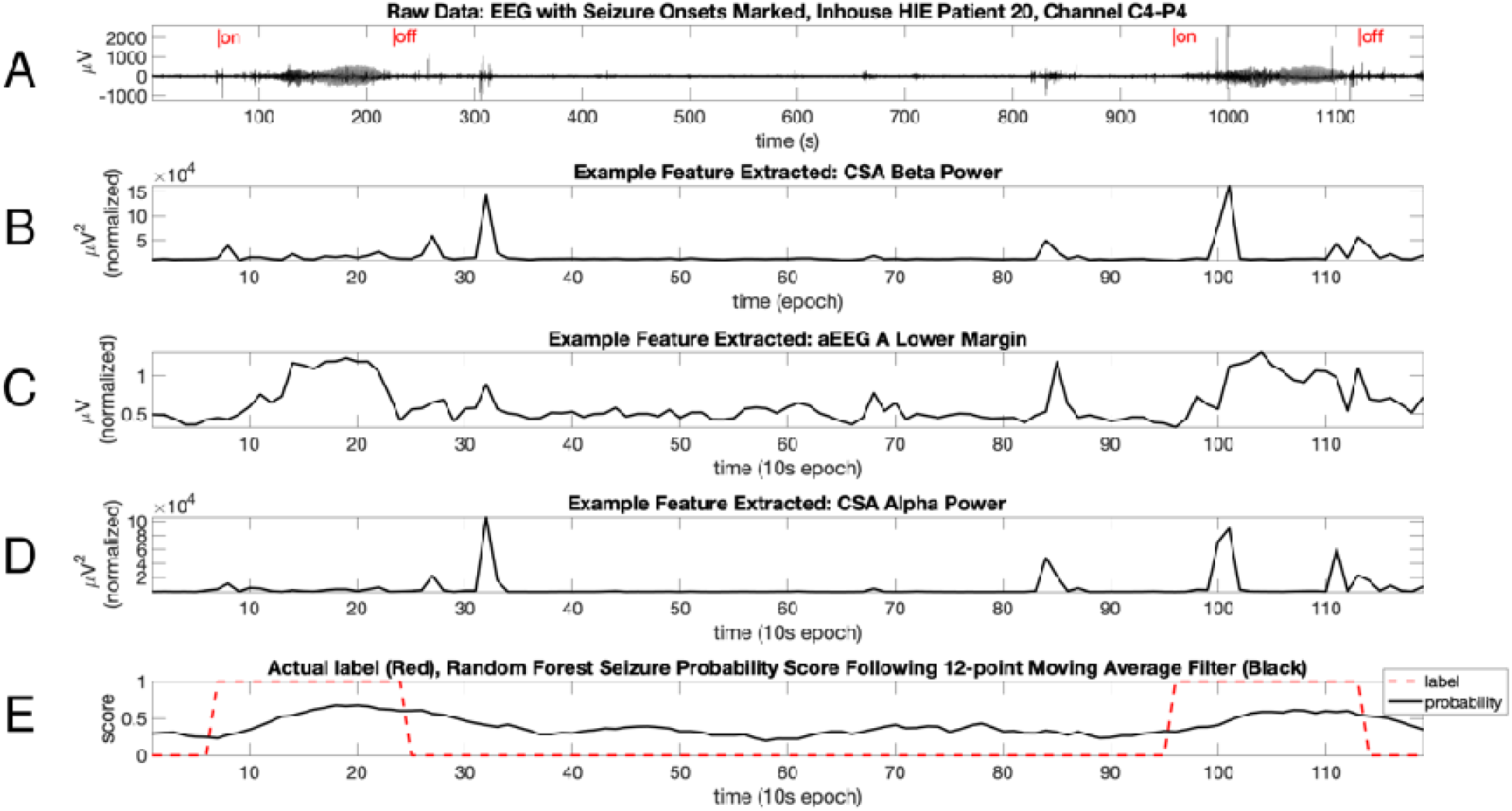
A. EEG from HIE Patient 20 from the inhouse testing. Channel C4-P4 plotted with EEG band-pass filtered from 0.5-40Hz. Seizure onsets and offsets marked using clinician label. Panels B, C, and D show the three features plotted for the same EEG epoch. Each features shows increases around seizure epoch. E. RF Classifier seizure probability scores following training and testing in black and true labels in red.

To generate the neonatal seizure detection algorithm, 11 clinical features were extracted from the two centroparietal electrode channels (C3-P3 and C4-P4). Each feature was computed across 10-second nonoverlapping EEG windows. Six aEEG clinical features were collected after computing a generic aEEG using the procedure adapted from Zhang and Ding et al 2013 (15), with 10-second windows used for feature extraction instead of the five minutes windows used in the original publication. These features are the aEEG upper margin, lower margin, and median envelope for two versions of the aEEG (aEEG A and aEEG B). These versions were generated by implementing different proprietary domain algorithm parameters extracted from studies published in the literature (Table 2) (16). For the CSA, five spectral features were collected. These features are the power across each of the Berger Bands (delta, theta, alpha, beta) and the spectral slope. The top three features previously found to have the highest importance are the CSA Beta power, aEEG lower margin, and CSA alpha power (Figure 1B, C, and D).

**Table 2:** Documented parameters for generating two versions of the aEEG.

| *aEEG*<br>*Version* | *Reference* | *Asymmetric Bandpass Filter (2-15Hz)* | *Enveloping Filter* |
| --- | --- | --- | --- |
| *aEEG A* | Werther et al 2018 | Linear Phase FIR filter (Parks-McClellan)<br>Filter 2 | Rectangular Moving Average Filter<br>(3-second window) |
| *aEEG B* | Werther et al 2018 | Linear Phase FIR filter (Parks-McClellan)<br>Filter 5 | Rectangular Moving Average Filter<br>(0.5-second window) |

### Artifact Removal and Data Normalization

Artifact removal was conducted using the extracted features. Epochs were removed if their aEEG lower margin values were less than 0.001µV or total power from 0.5-30Hz was less than 0.001µV^2^. Upper threshold artifacts were removed per patient. This process involved calculating the mean upper margin and mean total power from 0.5-30Hz across all the epochs of a given patient. Epochs were removed if their upper margin values or total power were greater than five times either of these mean values. Finally, scaling was also done per patient. Scaling was accomplished by subtracting each feature by its median and dividing by the interquartile range.

### Model Training, Testing, and Post-Processing

Random Forest (RF), Support Vector Machines (SVM), and feedforward Artificial Neural Network (ANN) classifiers were used for training and testing on MATLAB. To address the problem of class imbalance, synthetic seizure data was added to the training data using Adaptive Synthetic Sampling (17,18). The RF classifier was deployed using the MATLAB “fitcensemble” function with bagging method selected using 100 decision trees (19). To facilitate feature selection assessments, all 11 variables were included for sampling across every decision tree. The SVM was applied using the “fitcsvm” function using a radial basis function kernel and feature standardization was selected (20). The ANN was implemented using the “fitcnet” function with the default neural network architecture used (21). Z-score feature standardization was selected for using hyperparameter tuning.

Training and testing were done to evaluate algorithm performance. Training was done using all 79 patients in the external Helsinki dataset; of these patients, 39 had seizures. Testing was done on the 23 HIE holdout patients from the inhouse dataset of patients; of these patients, 10 had seizures. Two trained models were generated per classifier. One for the left electrode and the other for the right. Each model was tested and the maximum probability score across both models was used to evaluate algorithm performance.

### Model Post-Processing

All classifier probability scores were smoothed using a 12-epoch moving average window filter (Figure 1E). This window length is based on the average seizure duration of the training data which was 115 seconds in duration since each epoch in this study represented 10-seconds of EEG. It also falls within the range of the average neonatal seizure duration that spans anywhere between one to three minutes (22). ROC curves and patient-level accuracy were computed using the optimal patient-independent probability threshold for each classifier, derived with MATLAB’s “perfcurve” function. We then applied 1,000 bootstrap iterations to the test-set predictions to generate confidence intervals for all performance metrics.

## Results

### ROC Performance Assessments

The patient-independent performance of the algorithm was first tested using models trained on all 79 patients in the external dataset and tested on the 23 patients in the inhouse dataset. The area under the curve (AUC) for the RF, SVM, and ANN classifiers were 0.76, 0.77, and 0.77 respectively following AUC evaluation using 1000 bootstrap samples (Figure 2A).

**Figure 2.**
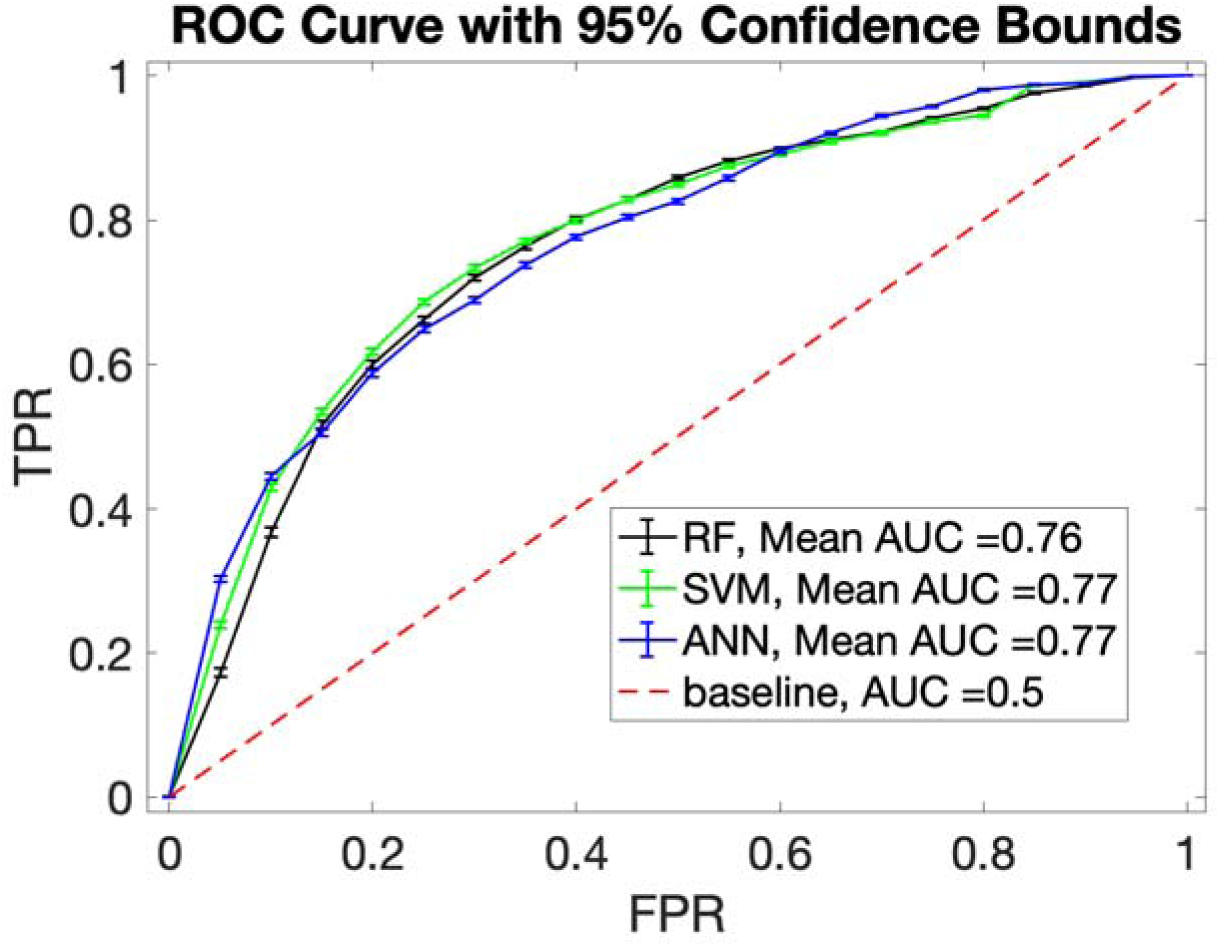
Patient-independent performances of aEEG and CSA detection algorithm using the RF, SVM, and ANN classifiers. A. Classifier performance on the testing dataset shows mean AUC scores of 0.76, 0.77 and 0.77 for RF, SVM and ANN respectively.

To determine the per patient performance, the distribution of patient accuracy scores was computed. Results were plotted in a histogram, and they show that the average patient accuracy scores were 0.85, 0.85, and 0.86 for the RF, SVM, and ANN respectively (Figure 3).

**Figure 3.**
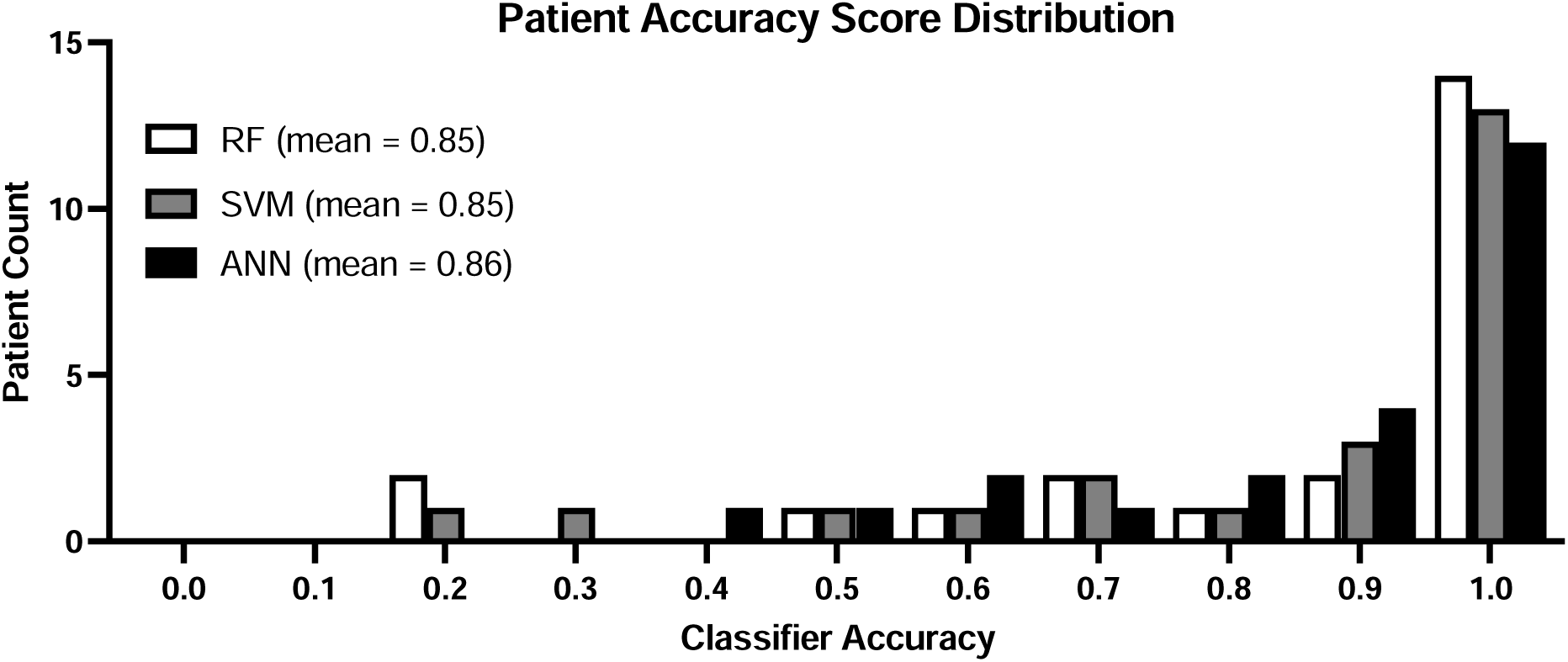
Distribution of accuracy scores for 23 HIE patients in the test set. Mean AUC score for RF, SVM and ANN classifiers are 0.85, 0.85, and 0.86 respectively.

### Algorithm Performance Assessments

#### Effect of Smoothing on Algorithm Performance

The effect of smoothing via moving average filter was quantified. This was done by assessing the RF classifier performance at seizure probability threshold of 0.5. These parameters were used in a previous study where we evaluated algorithm performance using patients of various etiologies. In this study, we focus on patients with only HIE. First, we assessed the effect of smoothing on false positive rate by plotting the false positive rate as a function of the moving average filter window length. We found that as the filter window increased, the false positive rate decreased (Figure 4A) going from 0.15 to 0.07. Next, we assessed the effect of smoothing on the percentage of seizures detected. When the decimal percentage of seizures detected is plotted as a function of filter window length, we see a clear decrease in seizure percentage detected with decimal percentages falling from 0.89 to 0.42 (Figure 4B). When seizure percentage and false positive rate are plotted against one another at discrete moving average window lengths, there is a clear direct relationship between increasing seizure percentage and increasing false positive rate (Figure 4C). Notably, at all window lengths, percentage of seizures detected exceeds the documented percentage of seizures detected by manual aEEG (12).

**Figure 4.**
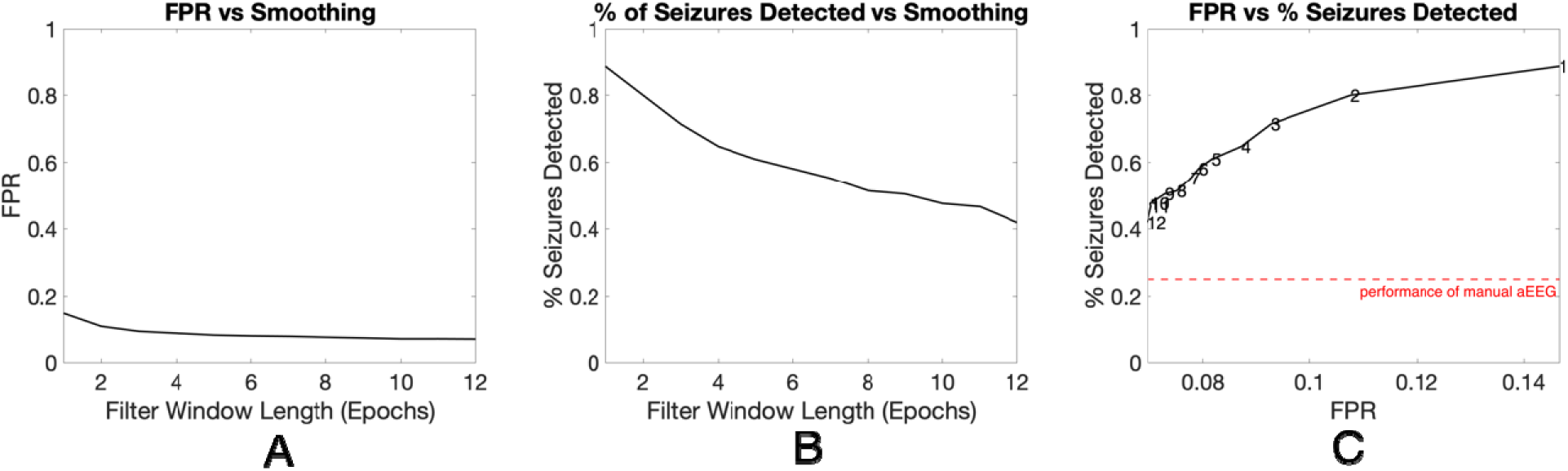
The effect of moving average filter on percentage of seizure detected and on false positive rate (FPR) using RF classifier at 0.5 seizure probability threshold. FPR decreases from 0.15 to 0.07 as filter window length. B. Decimal percentage of seizures detected drops from 0.89 to 0.42 as filter window increases. C. FPR plotted against percentage of seizures detected at discrete filter window lengths shows direct relationship between FPR and % of seizures detected. When compared to documented manual aEEG performance by Shellhaas et al 2007 algorithm performance at all window lengths exceeds manual aEEG performance (12).

#### Performance in Seizure-Positive HIE Patients and Analysis of Seizure Duration Effects

Only 10 neonates in our inhouse HIE cohort had seizures. We computed the average AUC of the 10 seizure patients per classifier, and found each classifier had a mean AUC of 0.85 with a median AUC of 0.84, 0.87, and 0.85 for the RF, SVM, and ANN classifiers respectively (Figure 5).

**Figure 5.**
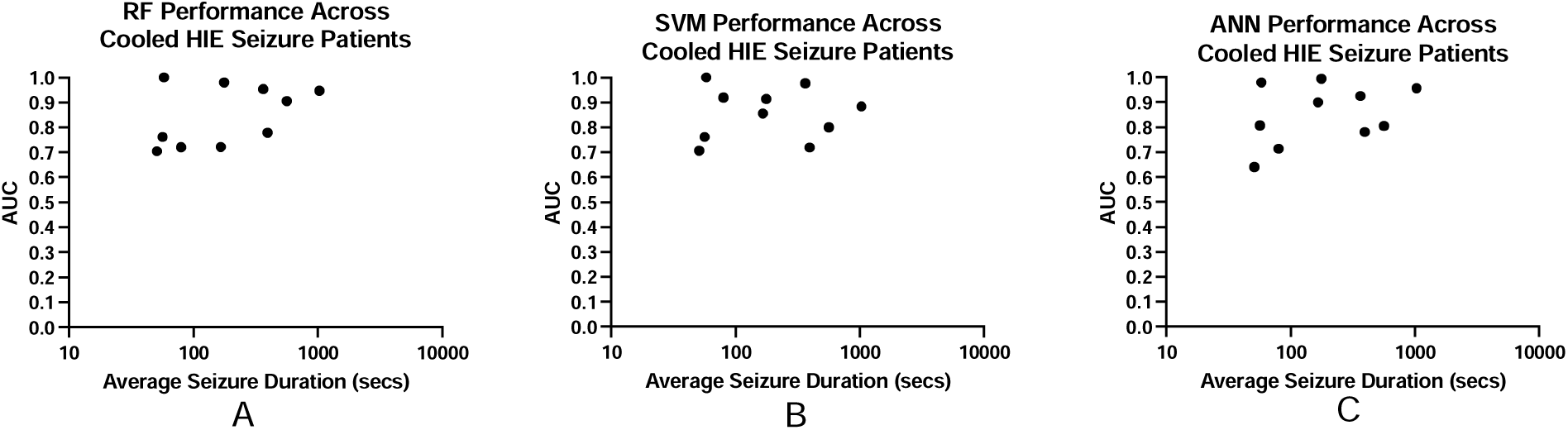
The effect of seizure duration on algorithm performance using AUC plotted as a function of average seizure duration of the 10 seizure patients. Seizure duration is not correlated with AUC for any of the classifiers based on Spearman correlation p-values. Also, the mean AUC per patient for all the classifiers is 0.85. Panel A. For RF, the P-value for Spearman correlation = 0.25. Panel B. For SVM, the P-value for Spearman correlation = 0.79. Panel C. For ANN, the P-value for Spearman correlation = 0.49.

Shorter duration seizures have been noted as more difficult to identify in manual aEEG (12,23). Thus, we aimed to see if our algorithm performed more poorly amongst HIE neonates with shorter average seizure duration. To do so, we plotted patient AUC values against average seizure duration for each classifier (Figure 5A-C). When we test the association between AUC values and seizure duration using Spearman correlation, we see there is no correlation for any of the three classifiers (p=0.25, 0.79, and 0.49 for RF, SVM, and ANN respectively).

## Discussion

### Previous Assessment of aEEG-CSA Ensemble Algorithm

This study validates an algorithm used for neonatal seizure detection on an independent cohort of 23 infants with HIE. This algorithm was initially assessed in a previous study using only patients in the external dataset via K-fold cross validation. The patient independent AUC reported in the previous study was 0.80, 0.69, and 0.79 for the RF, SVM, and ANN classifiers respectively. The previous study also showed that the algorithm had significantly lower accuracy across the 35 HIE patients compared to the other 44 patients with various etiologies. That finding motivated further assessment of algorithm performance on an independent HIE-only cohort.

### Potential Limitations Caused by Differences in Dataset

This study demonstrates that the aEEG-CSA algorithm detects seizures with similar sensitivity and specificity, with AUCs ranging from 0.76 to 0.77. This study used patients from a completely independent dataset from Helsinki University that included patients who were not explicitly cooled. Since our sample size is very limited (n=23), it is possible that performance can be improved by using a larger dataset of patients exclusively from the UChicago Comer hospital who also have similar disease etiology of HIE due to birth asphyxia.

### Effect of Moving Average Filter on Algorithm Performance

We found that increasing the filter window improved performance by lowering the false positive rate but with a tradeoff of lower seizure percentage detected. In both cases, the percentage of seizures detected remained well above 25% which is the documented manual performance in the largest study to date of aEEG performance on a per seizure basis from Shellhaas et al 2007 (12).

There, six EEG experts at varying levels of experience assessed 851 seizures across 121 patients. The future objective would be to combine smoothing with other computational methods for reducing the false positive rate such as a seizure onset or offset tolerance, so epochs directly before or after the seizure that might capture clinical seizure activity are not being flagged.

### No Correlation Between Algorithm Performance and Seizure Duration

In this cohort, there was no correlation between seizure duration and algorithm performance. Previous studies of manual aEEG performance have shown correlations between seizure duration and manual performance (12,24,25). The goal is for computer algorithms to help bridge the gap by improving seizure detection performance regardless of the patient seizure duration. Lack of correlation based on seizure duration and a mean AUC performance of 0.85 for seizure patients (Figure 5), shows that performance was consistently good across seizure patients regardless of their seizure duration.

## Conclusions

Using a cohort of 23 neonatal HIE patients with EEG recordings taken during cooling therapy, we found that the aEEG-CSA algorithm can detect seizures with a patient-independent AUC of 0.76, 0.77, and 0.77 for the RF, SVM and ANN classifiers respectively. The mean AUC for the 10 seizure patients was 0.85 for all three classifiers. Thus, seizure epochs in near-term HIE patients during cooling can still be detected even when training is done using a heterogeneous cohort of neonates with various disease etiologies. Finally, despite the tradeoffs of percentage of seizures detected and false positive rate when a moving average filter is used for postprocessing, algorithm performance still exceeds the standard manual aEEG’s performance for percentage of seizures detected.

## Data Availability

All data produced in the present study are available upon reasonable request to the authors

## Clinical Trial Number

Not applicable for this study.

## Declarations

### Ethics approval and consent to participate

This study was conducted in accordance with the World Medical Association Declaration of Helsinki. Because this was a retrospective analysis, written consent was not required.

### Consent for publication

All authors have provided consent for publication of this manuscript.

### Availability of data and materials

All data generated and analyzed in this study are included within this article. Additional inquiries may be directed to the corresponding author.

### Authors’ contributions

SE, JH, HD, and NPI conceptualized the study. SE performed aEEG feature extraction, and SE, NPI, and JH evaluated the aEEG features. SE conducted CSA feature extraction, and SE, NPI, and HD evaluated the CSA features. SE implemented the machine learning algorithms, and BBJ executed and evaluated the machine learning analyses. SE, NPI, and BBJ performed statistical analysis. SE and NPI drafted the manuscript, and SE, NPI, BBJ, and JH revised the manuscript. SE, NPI, BBJ, JH, and HD reviewed and approved the final version.

## Acknowledgments

We thank the University of Chicago Department of Pediatrics, Section of Neurology, the pediatric EEG technical staff at Comer Children’s Hospital, and the University of Chicago Department of Medicine for their support.

